# Knowledge and Positive Attitudes toward Caesarean Section Delivery among Married Women in Bangladesh

**DOI:** 10.64898/2026.04.02.26350021

**Authors:** Eusrat Jahan, Md. Mahir Faysal, Shuvashis Kumar Rimon

**Affiliations:** Department of Population Sciences, University of Dhaka, Dhaka-1000, Bangladesh

**Keywords:** Caesarean section, Knowledge, Attitude, Married women, Bangladesh

## Abstract

**Background:** Caesarean section (CS) rates in Bangladesh have increased rapidly in recent decades. This increase raises concerns about unnecessary procedures and their potential impacts on maternal health. Women’s knowledge and positive attitudes toward CS influence delivery preferences and decisions, yet these aspects remain underexplored in Bangladesh.

**Objectives:** To assess knowledge and positive attitudes toward CS and to identify factors associated with knowledge and positive attitudes among married women in Bangladesh.

**Methods:** The study utilized a cross-sectional sample of married women of reproductive age. A structured questionnaire was used in face-to-face interviews to collect data covering socio-demographic information, obstetric experiences, knowledge, and positive attitudes toward CS. Descriptive statistics, independent sample t-tests, and multiple linear regression analysis were performed to identify factors.

**Results:** This study showed that knowledge was lower among rural than urban women; lower among women with a previous CS than those without, and higher among women from husband-headed households. Additionally, respondents without an income source had higher knowledge than those with an income. Regarding attitudes, higher monthly family income was associated with more positive attitudes, while larger family size was associated with lower positive attitudes. Women in husband-headed households had more positive attitudes than those in other-headed households, and women with previous CS had lower positive attitudes. Importantly, higher knowledge scores were strongly associated with more positive attitudes toward CS.

**Conclusion:** Strengthening antenatal care, including health, educational, and counselling services, particularly for rural women, larger families, husband-headed households, and women with prior CS, could improve knowledge and promote informed, positive attitudes toward appropriate CS use. Policies and programs should prioritize rural outreach, improve provider–patient communication (especially after a CS), and ensure high-quality counselling, informed consent, and male-inclusive antenatal sessions to support the appropriate use of CS.

## Introduction

Caesarean section (CS) is a surgical procedure for delivering a baby when vaginal delivery becomes medically complicated and unsafe for the mother or newborn [1, 2]. The World Health Organization (WHO) recommends that national CS rates remain within 10%–15% of all births, as rates above this threshold are not associated with improved maternal or neonatal outcomes [3, 4]. Globally, CSs account for about 21% of all births and are projected to rise to 29% by 2030, indicating a continuing global increase in their use [5].

In Bangladesh, the prevalence of CS deliveries has increased dramatically over the past three decades-from 3% in 1990 to 18% in 2011, 24% in 2014, and 34% in 2017–18 [6, 7]. By 2022, the national CS rate reached 45%, far exceeding the WHO recommendation [8, 9]. Recent data suggest it rose further to 50.8% in 2023, representing one of the highest CS rates worldwide [9]. Thus, Bangladesh has experienced a significant increase in CS deliveries over the past two decades, highlighting an urgent national health concern.

While CS is lifesaving when medically indicated, the growing number of unnecessary CS deliveries presents serious implications for a developing country like Bangladesh [2]. Although CS can prevent adverse maternal and neonatal outcomes, it also increases the risk of postpartum hemorrhage, urinary complications, delayed fertility, psychological distress, and placental problems in future pregnancies [10–15]. Several non-clinical factors contribute to high CS rates, including profit-driven medical practices, misinformation, limited patient autonomy, and sociocultural influences [19–21]. Many women, particularly in rural areas, have a limited understanding of pregnancy risks and are often excluded from decisions regarding their delivery mode [22]. In slum areas, physicians frequently recommend CS without adequate counselling, leaving women unaware of the reasons for surgery [21]. Even without medical indication, repeat CSs or maternal-request procedures are common in private clinics [20].

Addressing the rising CS rate requires informed, patient-centered strategies. Enhancing women’s knowledge of the benefits and risks of different modes of childbirth can promote safe vaginal delivery and reduce elective CS [23, 24]. Empowered women are more likely to utilize maternal healthcare services and apply that information in choosing their delivery mode [24]. Structured antenatal education, midwife-led counselling, and community-based awareness programs may enable women to make informed decisions [23]. The economic burden is also substantial; CS delivery increases out-of-pocket costs, strains household finances, and misuses limited healthcare resources [16, 45]. High CS rates further stress hospital systems and heighten the risk of postnatal depression among mothers [17, 18]. Therefore, analyzing the most recent data to gain a better understanding of these patterns is necessary.

Although several studies in Bangladesh have explored socioeconomic and demographic determinants of CS [2, 14, 20, 25–37, 42–44] and aspects of the decision-making process [21, 38–41], few have examined women’s knowledge and attitudes toward CS using primary data. Existing research has largely focused on urban populations [23, 39] with limited evidence from rural settings [19, 22]. A comprehensive study by Yasmin et al. [22] examined knowledge, attitudes, and practices related to hospital deliveries in rural areas but did not specifically analyze factors influencing CS knowledge or attitudes. Consequently, a substantial research gap remains regarding how married women’s knowledge and attitudes affect the increasing trend of CS deliveries in Bangladesh.

Understanding these aspects is important for designing evidence-based interventions and policies that encourage safe and appropriate maternal healthcare practices. Therefore, this study analyses the knowledge and positive attitudes of married women toward caesarean section across both urban and rural areas of Bangladesh, thereby addressing the identified research gap and informing strategies to reduce unnecessary CS deliveries.

## Materials and Methods

### Participants and Procedure

The study employed a cross-sectional study design, where a questionnaire was created to collect information through in-person interviews conducted in the Mymensingh and Sylhet divisions, using purposive sampling. The urban area comprised three wards: Ward No. 8 (Panur), Ward No. 9 (Duttapara), and Ward No. 4 (Rawt Para) of Mohanganj Municipality in Netrokona District under Mymensingh Division. The rural data were collected from Horipur, Badehoripur, and Baghaucha villages of Joysree Union, located in Dharmapasha Thana, Sunamganj District, Sylhet Division.

As the prevalence rate was not found in existing literature, 0.5 was used in the equation to maximise the study’s sample size. The sample size was found to be 380 using the formula. However, due to limited time and resources, a total of 301 currently married women between 15 and 49 years of age in the reproductive period were interviewed. Out of these, 206 participants originated from rural regions, while the remaining 95 respondents were from urban areas, in accordance with the rural-urban population ratio observed in the 2022 Bangladesh Population & Housing Census [50]. A structured questionnaire in Bengali was employed to collect data during in-person interviews. The data were collected from both rural and urban areas, as well as from the respondents’ residences, allowing for a comprehensive and diverse range of perspectives to be captured from 25 January 2026 to 27 January 2026, after getting the oral concerns of the respondents. Those who experienced difficulty communicating or hearing, as well as those who were not in a suitable health condition to engage in conversation, were deliberately omitted from the study to ensure the validity and reliability of the data.

### Description of Variables

To measure the knowledge and positive attitudes toward CS among married women, a self-constructed Likert scale was used to measure opinions, attitudes, or behaviors [46]. The scale consists of 12 statements to assess knowledge and 10 statements to evaluate positive attitudes. The questions and the statements were modified according to the previous studies [47–49]. Responses were rated from 0 (strongly disagree) to 4 (strongly agree). Higher scores indicated higher knowledge and more positive attitudes. The scales showed good reliability, with Cronbach’s alpha values of 0.839 (knowledge) and 0.790 (attitudes).

The independent variables of interest in this study from existing literature review included respondents age, education, monthly income, place of residence, religion, having income source, age at marriage, use of mobile phone, use of internet, types of family, household head, number of family member, having any children, number of children, age of the first child, family monthly income, having a previous CS, perception of health, knowledge of CS and respondent’s husbands age, education, age at marriage, monthly income.

### Data Processing & Analysis

The Statistical Product and Service Solutions (SPSS) version 23 was used for data entry and analysis. Following data entry, frequencies were computed in SPSS to examine the distribution of socio-demographic variables at the univariate analysis level. Then, bivariate analysis was conducted to assess the relationship between the dependent and independent variables using an independent sample t-test when the independent variable had two categories and Pearson’s correlation when the variables were at an interval level. Later, the association between the dependent variable and multiple independent variables that were significant at the bivariate level was examined using multiple linear regression.

### Ethical Approval

The study was approved by the Academic Committee of the Department of Population Sciences, University of Dhaka (Approval No. DPSEB/A/004/2026). Before conducting the interviews, all respondents provided oral consent. Also, the study’s aim and participants’ right to withdraw at any point were explained. Data were stored securely on a password-protected personal computer to ensure confidentiality and privacy.

## Results

### Sample Characteristics

Table 1 shows the distribution of the respondents’ socio-demographic characteristics. Most respondents were aged between 15 to 30 years. Approximately two-thirds (68.4%) of the population lived in rural areas. and 83.7% of the respondents were Muslim. Only one-fourth (25.9%) reported having an income source, and among respondents with their own income, 55.1% had a monthly income of BDT 5000 or less. In terms of education, most respondents (82.4%) and their husbands (74.4%) had some level of education. Two-thirds of the respondents’ husbands were 30 or older (66.8%). Most women were married by the age of 18 or older (62.1%). Notably, about half (49.5%) of the respondents’ husbands earned ≤15,000 per month, and half (50.5%) earned more, while about 55.5% of families reported a monthly income above 15,000. Among the respondents, mobile phone ownership was high (91.7%), but internet use was lower (35.5%). More than two-thirds of households were nuclear families (68.4%), and the majority of households were headed by the participant’s husband (68.1%). The majority had at least one child (86.7%). Lastly, a previous CS was reported by around two-thirds (64.5%) of the respondents.

**Table 1:** Respondents’ socio-demographic characteristics

| Characteristics | N | % |
| --- | --- | --- |
| **Age of the respondents** |  |  |
| 15-30 | 185 | 61.5 |
| 30+ | 116 | 38.5 |
| **Place of residence of the respondents** |  |  |
| Urban | 95 | 31.6 |
| Rural | 206 | 68.4 |
| **Religion of the respondents** |  |  |
| Muslim | 252 | 83.7 |
| Hindu | 49 | 16.3 |
| **Having an income source of the respondents** |  |  |
| Yes | 78 | 25.9 |
| No | 223 | 74.1 |
| **Monthly income of the respondents** |  |  |
| ≤ 5000 | 43 | 55.1 |
| 5000+ | 35 | 44.9 |
| **Education of the respondents** |  |  |
| No schooling | 53 | 17.6 |
| Any schooling | 248 | 82.4 |
| **Age of the respondents' husbands** |  |  |
| ≤30 | 100 | 33.2 |
| 30+ | 201 | 66.8 |
| **Education of the respondents' husbands** |  |  |
| No schooling | 77 | 25.6 |
| Any schooling | 224 | 74.4 |
| **Age at marriage of the respondents** |  |  |
| Below 18 | 114 | 37.9 |
| 18+ | 187 | 62.1 |
| **Monthly income of the respondents' husbands** |  |  |
| ≤15000 | 149 | 49.5 |
| 15000+ | 152 | 50.5 |
| **Monthly income of the respondents' family** |  |  |
| ≤15000 | 134 | 44.5 |
| 15000+ | 167 | 55.5 |
| **Use of mobile phone** |  |  |
| Yes | 276 | 91.7 |
| No | 25 | 8.3 |
| **Use of the internet** |  |  |
| Yes | 98 | 35.5 |
| No | 178 | 64.5 |
| **Types of family** |  |  |
| Nuclear | 206 | 68.4 |
| Joint | 95 | 31.6 |
| **Household head** |  |  |
| Other members of the family | 96 | 31.9 |
| Husband of the respondents | 205 | 68.1 |
| **Having any children** |  |  |
| Yes | 261 | 86.7 |
| No | 40 | 13.3 |
| **Having a previous CS** |  |  |
| Yes | 194 | 64.5 |
| Others | 107 | 35.5 |

### Knowledge regarding CS delivery among married women

Table 2 shows knowledge regarding CS among married women in Bangladesh. Most of the respondents were aware of CS (77.4%). Nearly one-third recognized infection risks (31.6%), while many were uncertain (29.2%) about their impact on future pregnancies. Notably, over half (58.1%) strongly disagreed that normal delivery is possible after CS. For clinical indications, about one-fourth strongly agreed that CS is necessary for a large baby (28.6%) or delayed labor (26.9%), whereas 30.2% strongly disagreed with its necessity for an inverted baby. Regarding cost and care, more than half (52.8%) strongly agreed that CS is more expensive than normal delivery, and most reported the need for rest and an extended hospital stay after CS.

**Table 2:**
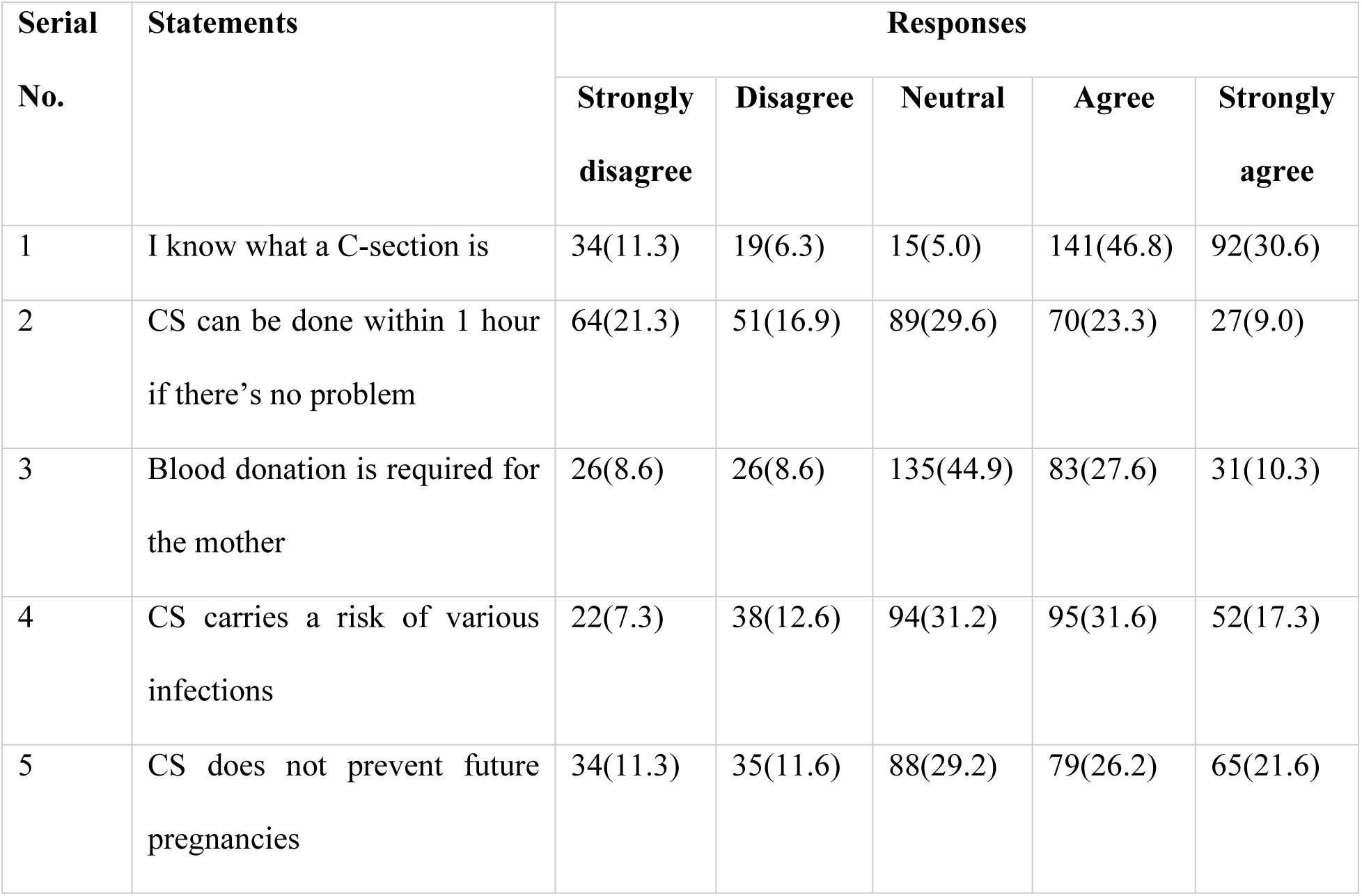
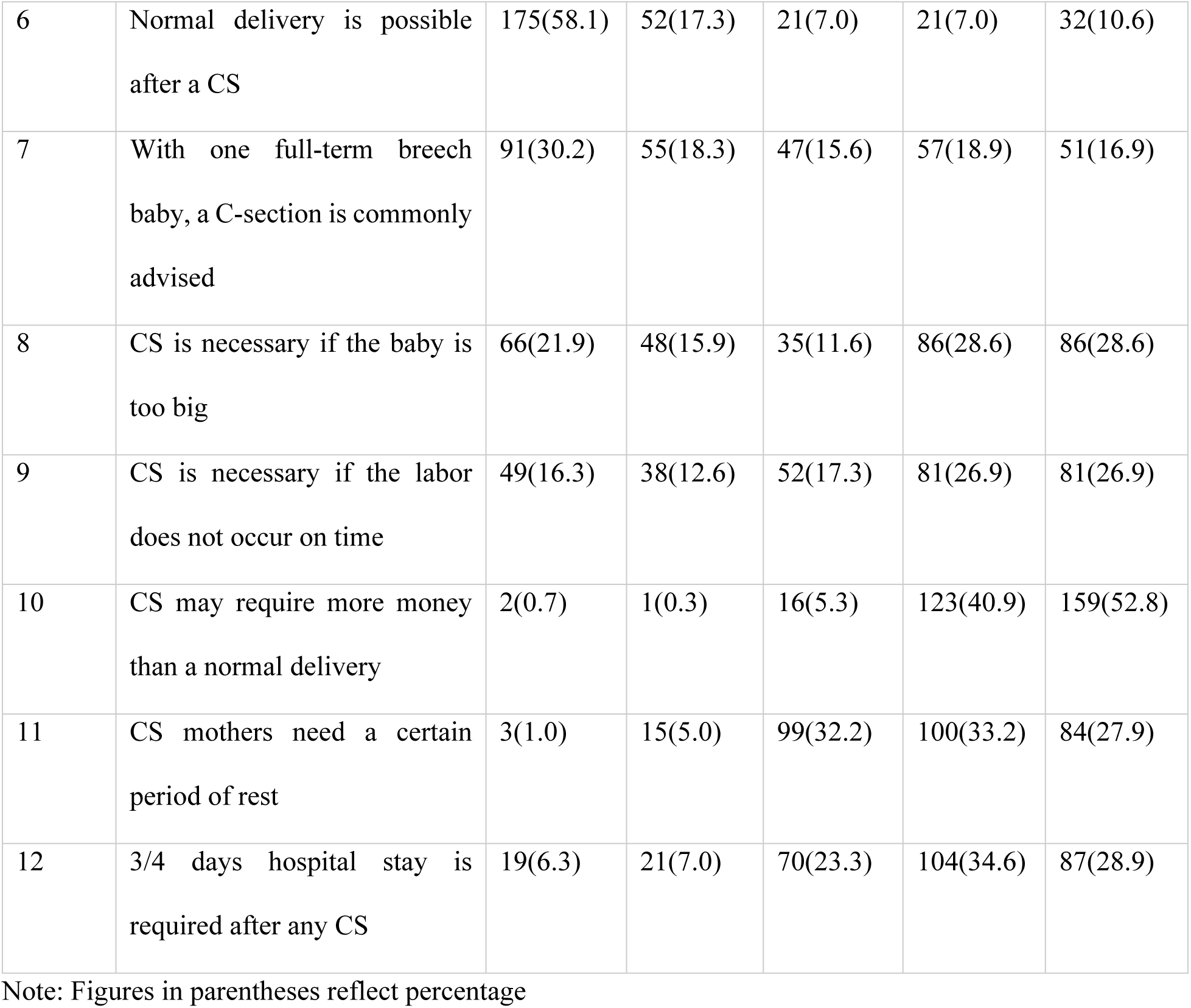
Distribution of knowledge regarding CS delivery

| Serial No. | Statements | Responses |  |  |  |  |
| --- | --- | --- | --- | --- | --- | --- |
|  |  | Strongly disagree | Disagree | Neutral | Agree | Strongly agree |
| 1 | I know what a C-section is | 34(11.3) | 19(6.3) | 15(5.0) | 141(46.8) | 92(30.6) |
| 2 | CS can be done within 1 hour if there's no problem | 64(21.3) | 51(16.9) | 89(29.6) | 70(23.3) | 27(9.0) |
| 3 | Blood donation is required for the mother | 26(8.6) | 26(8.6) | 135(44.9) | 83(27.6) | 31(10.3) |
| 4 | CS carries a risk of various infections | 22(7.3) | 38(12.6) | 94(31.2) | 95(31.6) | 52(17.3) |
| 5 | CS does not prevent future pregnancies | 34(11.3) | 35(11.6) | 88(29.2) | 79(26.2) | 65(21.6) |
| 6 | Normal delivery is possible after a CS | 175(58.1) | 52(17.3) | 21(7.0) | 21(7.0) | 32(10.6) |
| 7 | With one full-term breech baby, a C-section is commonly advised | 91(30.2) | 55(18.3) | 47(15.6) | 57(18.9) | 51(16.9) |
| 8 | CS is necessary if the baby is too big | 66(21.9) | 48(15.9) | 35(11.6) | 86(28.6) | 86(28.6) |
| 9 | CS is necessary if the labor does not occur on time | 49(16.3) | 38(12.6) | 52(17.3) | 81(26.9) | 81(26.9) |
| 10 | CS may require more money than a normal delivery | 2(0.7) | 1(0.3) | 16(5.3) | 123(40.9) | 159(52.8) |
| 11 | CS mothers need a certain period of rest | 3(1.0) | 15(5.0) | 99(32.2) | 100(33.2) | 84(27.9) |
| 12 | 3/4 days hospital stay is required after any CS | 19(6.3) | 21(7.0) | 70(23.3) | 104(34.6) | 87(28.9) |

Note: Figures in parentheses reflect percentage

### Correlates of Knowledge regarding CS in Bangladesh

Table 3 shows that knowledge regarding CS had a negative association with the number of family members (r = -0.114, p = 0.049). On the other hand, respondents’ monthly income, years of schooling, age at marriage, number of children, age of first child, perception of health, family’s monthly income, and respondents’ husbands’ age, years of schooling, age at marriage, and monthly income were not found to be statistically significantly correlated.

**Table 3:** Correlation of Knowledge regarding CS with socio-economic variables

| **Variables** | **Pearson r** | **p-value** |
| --- | --- | --- |
| Age of the respondents | 0.057 | 0.327 |
| Monthly income of the respondents | 0.185 | 0.105 |
| Years of schooling of the respondents | 0.058 | 0.316 |
| Age of the respondents' husbands | 0.038 | 0.509 |
| Years of schooling of the respondents' husbands | -0.016 | 0.788 |
| Age at marriage of the respondents | -0.044 | 0.451 |
| Age at marriage of the respondents' husbands | -0.089 | 0.124 |
| Monthly income of the respondents' husbands | 0.083 | 0.153 |
| Monthly income of the respondents' family | 0.031 | 0.589 |
| Number of family members | -0.114 | 0.049 |
| Number of children | -0.072 | 0.243 |
| Age of the first child | 0.095 | 0.124 |
| Perception of health of the respondents | 0.009 | 0.870 |

Note: Correlation is significant at the 0.05 level (2-tailed)

Table 4 shows that the mean of knowledge regarding CS among married women was significantly greater in urban (33.28) than rural settings (25.28). Respondents without a source of income reported a higher knowledge (28.44) compared to those with an income source (25.97). The knowledge was also higher among respondents from husband-headed households (28.63) than those from other types of households (26.04). Lastly, respondents with a previous CS delivery reported a lower knowledge (26.84) compared to those without a previous CS delivery (29.55). All of these differences showed statistical significance (p < 0.05).

**Table 4:** Independent sample t-test of knowledge regarding CS among married women

| Category | N | Mean | Std. Deviation | p-value |
| --- | --- | --- | --- | --- |
| **Place of residence** |  |  |  | <0.001 |
| Urban | 95 | 33.28 | 7.68 |  |
| Rural | 206 | 25.28 | 8.23 |  |
| **Religion** |  |  |  | 0.653 |
| Islam | 252 | 27.70 | 9.09 |  |
| Others | 49 | 28.33 | 7.68 |  |
| **Having an income source** |  |  |  | 0.034 |
| No | 223 | 28.44 | 8.56 |  |
| Yes | 78 | 25.97 | 9.51 |  |
| **Use of mobile phone** |  |  |  | 0.425 |
| No | 25 | 29.16 | 8.25 |  |
| Yes | 276 | 27.68 | 8.93 |  |
| **Use of the internet** |  |  |  | 0.265 |
| No | 178 | 27.24 | 9.18 |  |
| Yes | 98 | 28.49 | 8.43 |  |
| **Types of family** |  |  |  | 0.830 |
| Joint | 95 | 27.64 | 8.95 |  |
| Nuclear | 206 | 27.88 | 8.85 |  |
| **Household head** |  |  |  | 0.018 |
| Other members of the family | 96 | 26.04 | 9.28 |  |
| Husband | 205 | 28.63 | 8.56 |  |

| Having any children |  |  |  | 0.338 |
| --- | --- | --- | --- | --- |
| No | 40 | 26.55 | 10.20 |  |
| Yes | 261 | 28.00 | 8.65 |  |
| Having a previous CS |  |  |  | 0.011 |
| Others | 107 | 29.55 | 8.82 |  |
| Yes | 194 | 26.84 | 8.77 |  |

Note: p-value indicates significance at the 95% level.

Table 5 shows the findings of the regression model analysis of the correlates associated with knowledge regarding CS among married women. Only the independent variables that showed significance in the bivariate analysis were included in the regression model after verifying the assumptions and checking for multicollinearity. The table shows that place of residence was a strong predictor. Residing in a rural area was related to a 7.437 point lower knowledge score than residing in an urban area (B=-7.437, p<0.001). Respondents with an income source had lower knowledge than those without (B= -2.420, p = 0.022). Additionally, respondents living in households headed by husbands showed higher knowledge (B= 2.479, p = 0.023) compared to those in households headed by others. Finally, a previous CS delivery was associated with lower knowledge (B= -2.901, p = 0.005) compared to those without a previous CS delivery. Overall, the model explains 20.9% of the variance in the dependent variable (adjusted R-squared = 0.209) and is statistically significant.

**Table 5:**
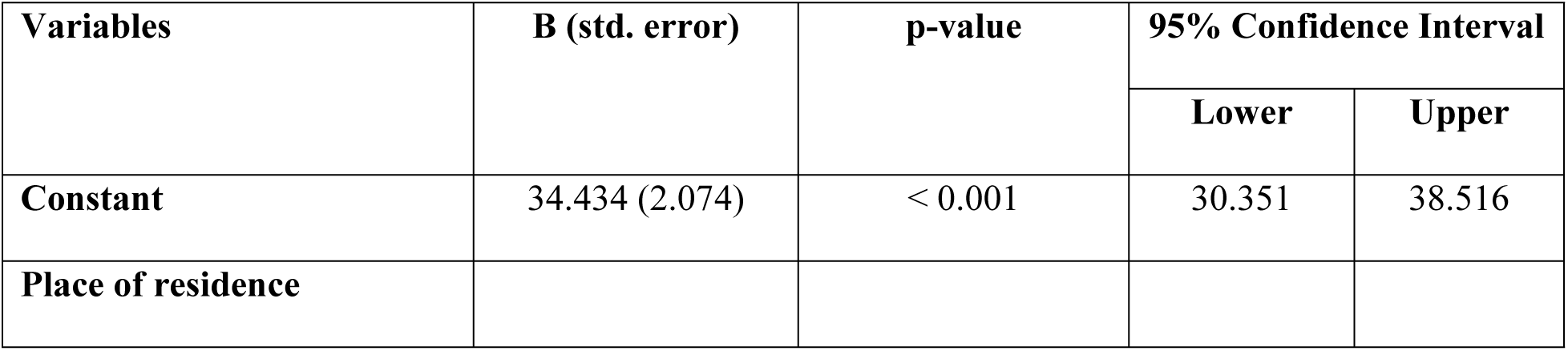
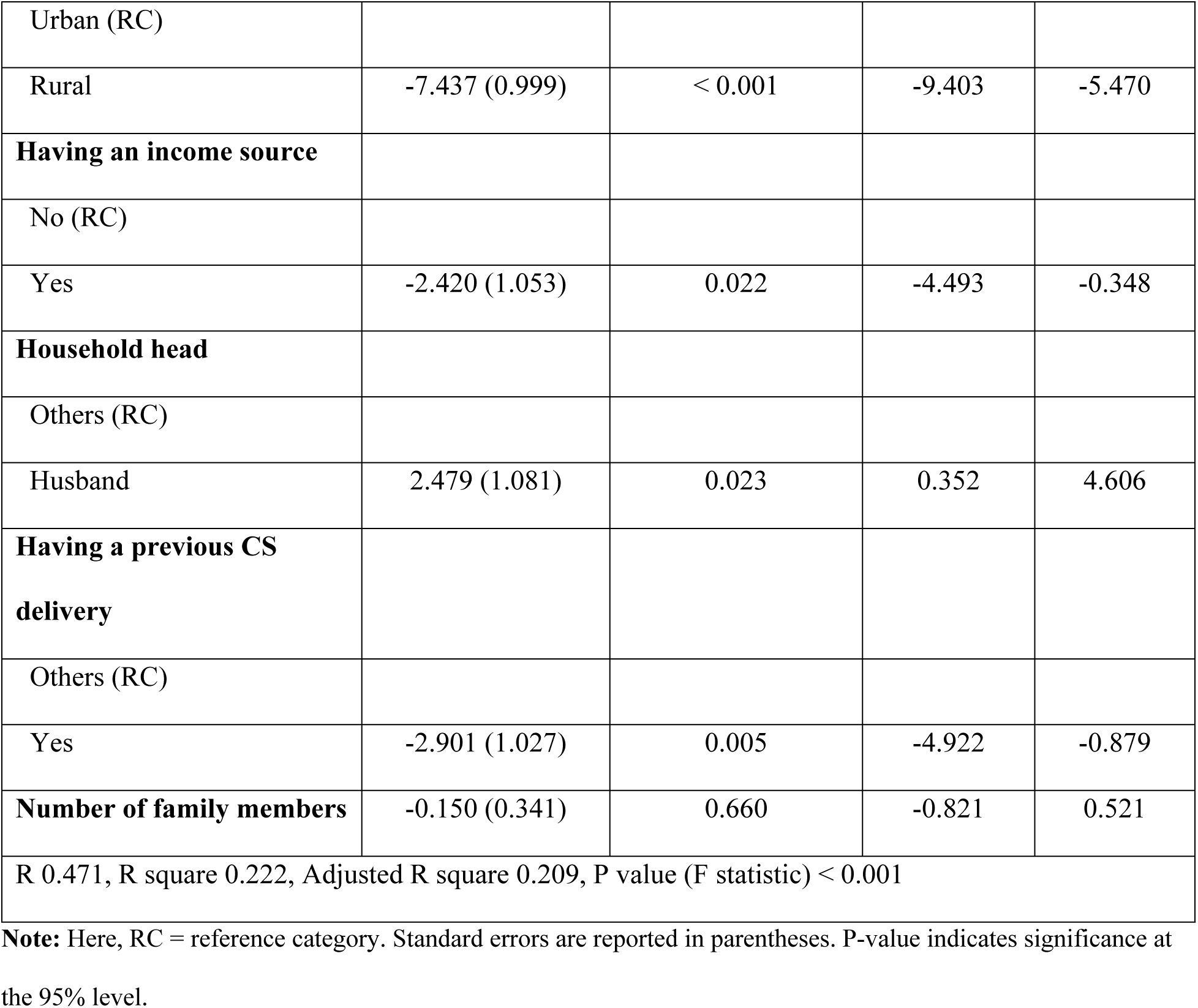
Regression results of knowledge regarding CS among married women

### Positive Attitudes toward CS delivery among married women

Table 6 shows the positive attitudes toward CS among married women in Bangladesh. Most respondents strongly disagreed that CS can be done without a doctor’s advice (52.8%) and that they were interested in having a CS (40.5%), while more than one-third (37.9%) strongly agreed that preparation can be done in advance. However, views on benefits were mixed. For example, 33.6% agreed that CS avoids health risks for mother and baby, while 32.2% strongly disagreed that CS is safer than normal delivery. Moreover, about one-third of respondents (30.6%) strongly disagreed that a CS baby has a stronger immune system. Lastly, 30.9% respondents agreed that mothers without health risks can also undergo CS.

**Table 6:** Distribution of positive attitudes toward CS delivery

| Serial No. | Statements | Responses |  |  |  |  |
| --- | --- | --- | --- | --- | --- | --- |
|  |  | Strongly disagree | Disagree | Neutral | Agree | Strongly agree |
| 1 | Interested in having CS | 122(40.5) | 41(13.6) | 36(12.0) | 38(12.6) | 38(12.6) |
| 2 | CS is safer than normal delivery | 97(32.2) | 52(17.3) | 79(26.2) | 38(12.6) | 35(11.6) |
| 3 | CS is less painful | 60(19.9) | 59(19.6) | 36(12.0) | 52(17.3) | 94(31.2) |
| 4 | Baby's due date can be chosen in CS | 47(15.6) | 34(11.3) | 65(21.6) | 91(30.2) | 64(21.3) |
| 5 | Preparation can be done in advance | 39(13.0) | 20(6.6) | 30(10.0) | 98(32.6) | 114(37.9) |
| 6 | CS avoids health risks for mother and baby | 16(5.3) | 38(12.6) | 97(32.2) | 101(33.6) | 49(16.3) |
| 7 | CS baby has a stronger immune system | 92(30.6) | 55(18.3) | 69(22.9) | 51(16.9) | 34(11.3) |
| 8 | CS can be done without a doctor's advice | 159(52.8) | 83(27.6) | 44(14.6) | 7(2.3) | 8(2.7) |
| 9 | CS babies have faster mental development | 16(5.3) | 29(9.6) | 111(36.9) | 103(34.2) | 42(14.0) |
| 10 | Mothers without health risks can also undergo CS | 57(18.9) | 27(9.0) | 41(13.6) | 93(30.9) | 83(27.6) |

Note: Figures in parentheses reflect percentage

### Correlates of Positive Attitudes toward CS in Bangladesh

Table 7 shows that the positive attitudes toward CS had a statistically significant negative correlation with age of the respondents (r=-0.255, p<0.001), age of the respondents’ husband (r=-0.251, p<0.001), number of family member (r=-0.288, p<0.001), number of children (r=-0.412, p<0.001), age of first child (r=-0.351, p<0.001). On the other hand, the attitudes toward CS had a statistically significant positive correlation with respondents’ monthly income (r=0.434, p<0.001), education (r=0.378, p<0.001), age at marriage (r=0.256, p<0.001), family’s monthly income (r=0.386, p<0.001), perception of health (r=0.297, p<0.001), knowledge of CS (r=0.203, p<0.001) and respondents’ husbands’ education (r=0.346, p<0.001), monthly income (r=0.303, p<0.001).

**Table 7:**
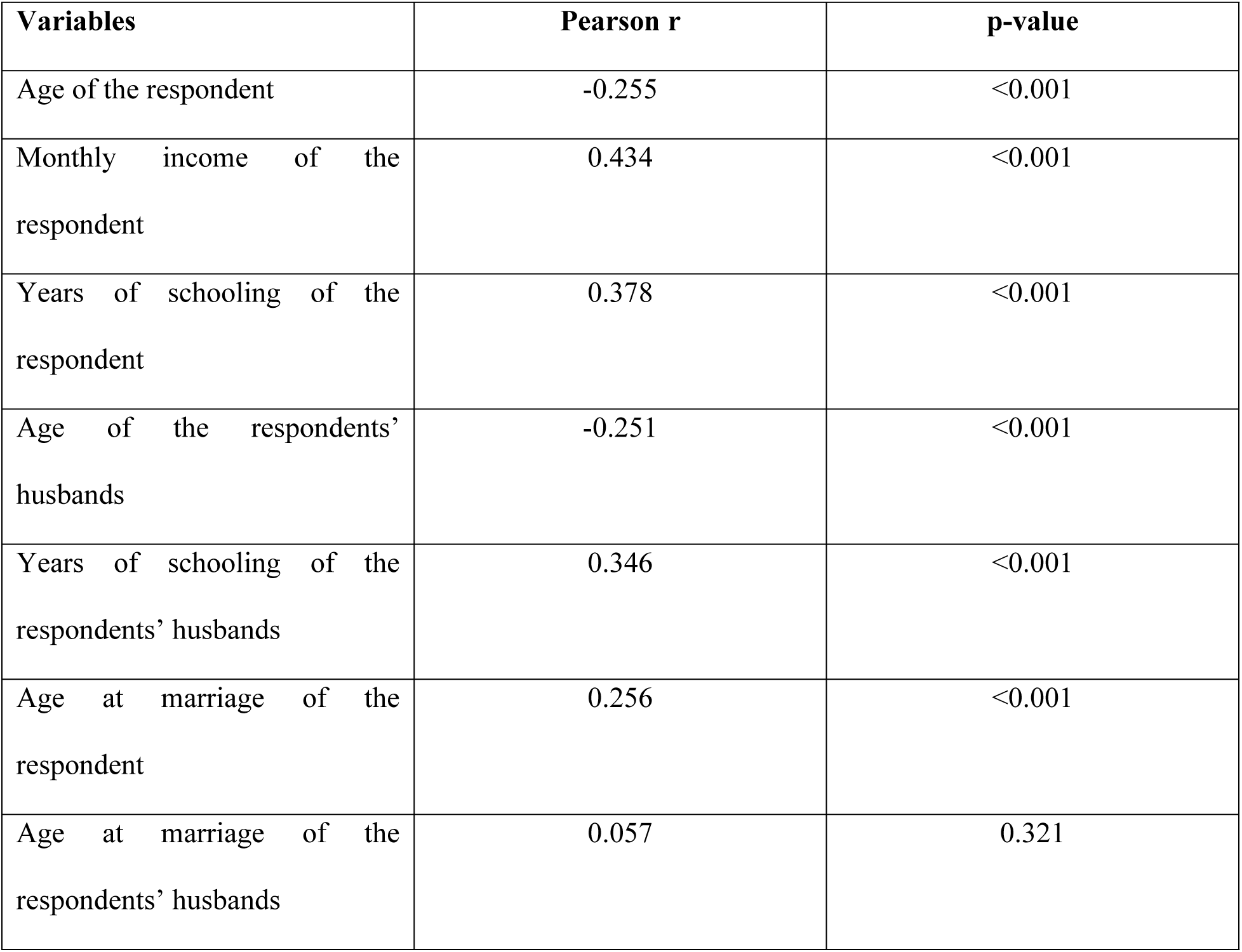
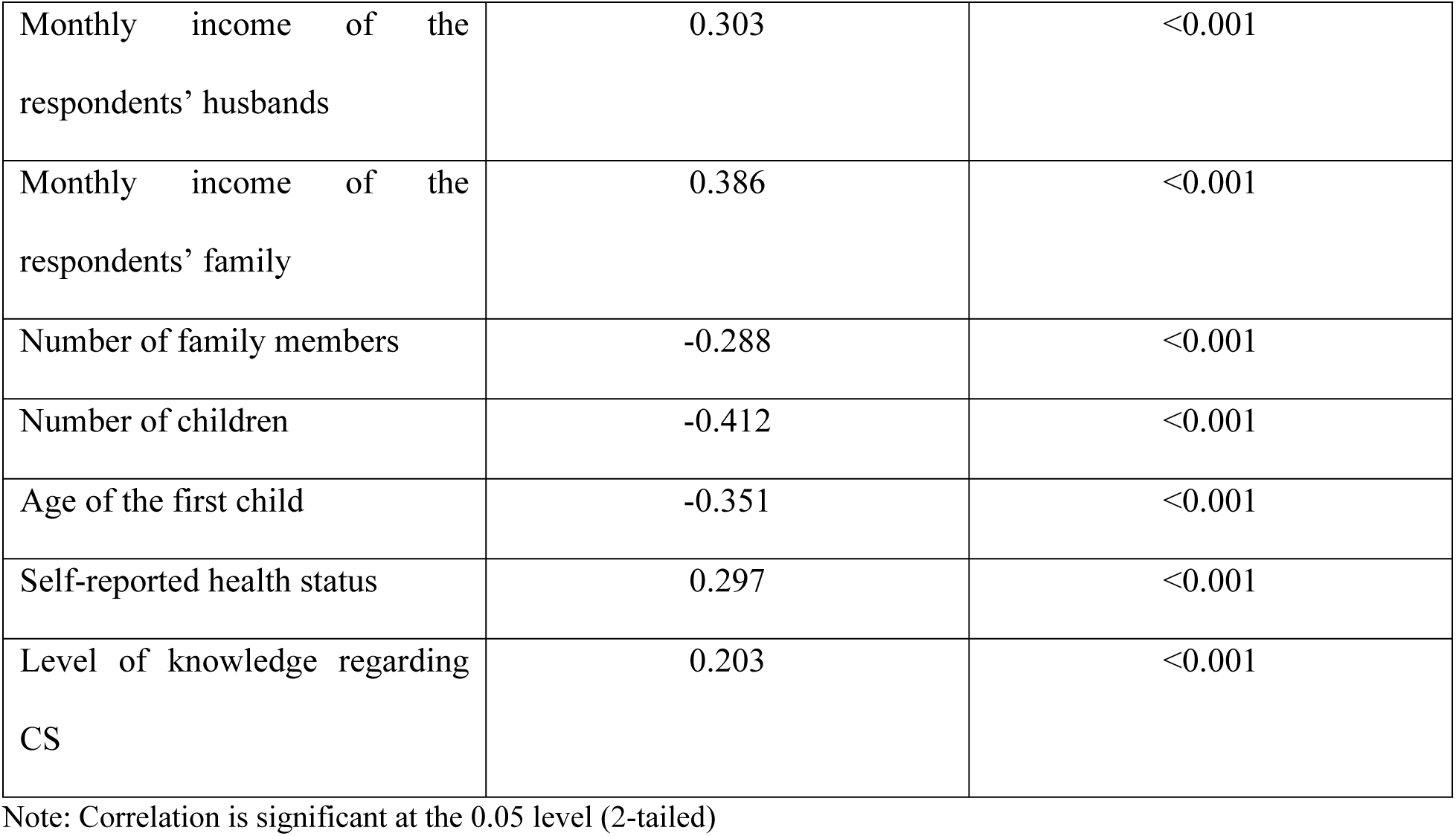
Correlation of positive attitudes toward CS with socio-economic variables

Table 8 shows that the mean of positive attitudes toward CS among married women was significantly lower in rural areas (19.05) than in urban areas (21.77). Muslim women had a lower positive attitude (19.10) than women in other religions (24.06). Respondents with a source of income reported a higher positive attitude (22.50) compared to those without an income source (19.00). Mobile phone users showed higher positive attitudes (20.49) compared to non-users (13.48). Internet users also had higher positive attitudes (24.31) than non-users (18.39). Respondents who lived in husband-headed households showed lower positive attitudes (18.54) than those who lived in households headed by someone other (22.83). Women without children had higher positive attitudes (22.60) than women with children (19.49). Finally, women who had not had a previous CS showed higher positive attitudes (25.12) than those who had (17.03). All of these differences showed statistical significance (p < 0.05).

**Table 8:** Independent sample t-test of positive attitudes toward CS among married women

| Category | N | Mean | Std. Deviation | p-value |
| --- | --- | --- | --- | --- |
| **Place of residence** |  |  |  | 0.004 |
| Urban | 95 | 21.77 | 6.72 |  |
| Rural | 206 | 19.05 | 8.02 |  |
| **Religion** |  |  |  | <0.001 |
| Islam | 252 | 19.10 | 7.54 |  |
| Others | 49 | 24.06 | 7.38 |  |
| **Having an income source of the respondent** |  |  |  | 0.001 |
| No | 223 | 19.00 | 7.61 |  |
| Yes | 78 | 22.50 | 7.50 |  |
| **Use of mobile phone** |  |  |  | <0.001 |
| No | 25 | 13.48 | 6.90 |  |
| Yes | 276 | 20.49 | 7.54 |  |
| **Use of the internet** |  |  |  | <0.001 |
| No | 178 | 18.39 | 7.19 |  |
| Yes | 98 | 24.31 | 6.64 |  |
| **Types of family** |  |  |  | 0.056 |
| Joint | 95 | 21.16 | 7.67 |  |
| Nuclear | 206 | 19.33 | 7.70 |  |
| **Household head** |  |  |  | <0.001 |
| Other members of the family | 96 | 22.83 | 7.21 |  |
| Husband | 205 | 18.54 | 7.59 |  |
| **Having any children** |  |  |  |  |
| No | 40 | 22.60 | 6.45 | 0.018 |
| Yes | 261 | 19.49 | 7.83 |  |
| **Having a previous CS** |  |  |  | <0.001 |
| Others | 107 | 25.12 | 6.58 |  |
| Yes | 194 | 17.03 | 6.75 |  |

Table 9 shows the findings of the regression model analysis of the correlates for positive attitudes toward CS among married women. The independent variables that showed significance at the bivariate level were entered in the regression model after checking assumptions and multicollinearity. The table indicates a positive association between the monthly income of families and positive attitudes (B= 0.00005185, p = 0.018). On the other hand, the number of family members was negatively associated with positive attitudes (B= -1.031, p = 0.009); each additional family member was associated with a 1.031-point decrease in positive attitudes scores. Households headed by the husband had lower positive attitudes compared to other-headed households (B= -2.724, p = 0.007). Having a previous CS delivery was associated with a substantially lower positive attitude than without a previous CS delivery (B= -5.933, p < 0.001); women with a prior CS had 5.933 points lower positive attitudes than those without. Lastly, a higher knowledge scale was associated with more positive attitudes (B= 0.112, p = 0.030); every one-unit increase in knowledge was related to a 0.112-point increase in positive attitudes. Overall, the model explains 42.5% of the variance in the dependent variable (adjusted R-squared = 0.425) and is statistically significant.

**Table 9:** Regression results of positive attitudes toward CS among married women

| Variables | B (std. error) | p-value | 95% Confidence Interval |  |
| --- | --- | --- | --- | --- |
|  |  |  | Lower | Upper |
| **Constant** | 25.385 (4.876) | < 0.001 | 15.776 | 34.993 |
| **Age of the respondent** | -0.121 (0.080) | 0.133 | -0.279 | 0.037 |
| **Education of the respondent** | -0.032 (0.114) | 0.778 | -0.258 | 0.193 |
| **Education of the respondents' husbands** | 0.045 (0.090) | 0.617 | -0.132 | 0.223 |
| **Age at marriage of the respondent** | 0.328 (0.191) | 0.086 | -0.047 | 0.704 |
| **Monthly income of the respondents' family** | $5.19 \times 10^{-5}$ ($1.00 \times 10^{-5}$) | 0.018 | 0.000 | 0.000 |
| **Number of family members** | -1.031 (0.391) | 0.009 | -1.803 | -0.260 |
| **Number of children** | -0.294 (0.605) | 0.628 | -1.486 | 0.898 |
| **Perception of health** | 0.021 (0.329) | 0.949 | -0.628 | 0.670 |
| **Place of residence of the respondent** |  |  |  |  |
| Urban (RC) |  |  |  |  |
| Rural | -0.906 (0.978) | 0.355 | -2.833 | 1.022 |
| **Religion of the respondent** |  |  |  |  |
| Islam (RC) |  |  |  |  |
| Others | -0.605 (1.121) | 0.590 | -2.815 | 1.604 |
| **Having an income source of the respondent** |  |  |  |  |
| No (RC) |  |  |  |  |
| Yes | 1.703 (0.918) | 0.065 | -0.107 | 3.513 |
| **Using internet** |  |  |  |  |
| No (RC) |  |  |  |  |
| Yes | 0.938 (1.129) | 0.407 | -1.288 | 3.164 |
| **Household head** |  |  |  |  |
| Others (RC) |  |  |  |  |
| Husband | -2.724 (1.008) | 0.007 | -4.711 | -0.737 |
| **Having a previous CS delivery** |  |  |  |  |
| Others (RC) |  |  |  |  |
| Yes | -5.933 (0.953) | < 0.001 | -7.811 | -4.055 |
| **Level of Knowledge of the respondent regarding CS** | 0.112 (0.051) | 0.030 | 0.011 | 0.212 |
| R 0.679, R square 0.461, Adjusted R square 0.425, P value (F statistic) < 0.001 |  |  |  |  |

**Note:** Here, RC = reference category. Standard errors are reported in parentheses. P-value indicates significance at the 95% level.

## Discussion

This study aimed to assess both knowledge and positive attitudes toward CS delivery in Bangladesh, with a focus on married women in both rural and urban areas. By analyzing various socio-demographic factors from both urban and rural areas, the research provides timely insights and a comprehensive evaluation to understand the knowledge and positive attitudes that are crucial for developing targeted interventions and policies, considering the increasing rate of unnecessary CS deliveries in Bangladesh.

The present study revealed several statistically significant findings related to knowledge of CS delivery. The study found that rural women had lower knowledge than urban women. This may be because urban women are more likely to own mobile phones and have access to technology, providing them with greater information [51]. This could be linked to previous studies, showing higher CS rates in urban than rural areas [16, 31, 32, 44, 52]. A further plausible explanation is that urban women have greater access to counseling and information from healthcare providers in both public and private facilities. In contrast, rural women face barriers such as limited-service availability, poor transportation, and communication problems [53, 54].

The study also found that respondents without an income source reported a higher knowledge compared to those with an income source. A possible explanation is that working women have less free time to attend community awareness programs, watch TV, use social media, or discuss health-related information with peers. Another possible reason could be that most respondents were engaged in low-skill jobs, which require long hours and leave little time for interacting with their neighbors or peers, learning, and seeking health-related information, leading to lower knowledge. Again, working women may have less time for postoperative CS recovery due to work demands. Moreover, their restricted time can even lead them to skip antenatal care and may encourage a preference for normal delivery. This aligns with previous findings of different studies that working women were less likely to undergo CS delivery than non-working women due to these reasons [29, 35, 55,56].

The study revealed that respondents from husband-headed households reported higher knowledge than those from female or other-headed households. This may be consistent with greater maternal healthcare use in male-headed households [57] and their relatively higher wealth [58, 59]. This may reflect women’s limited employment and unpaid domestic responsibilities, which reduce their financial capacity for healthcare and learning about CS. Further, husband-led households may provide more spousal support or access to health information by maternal health knowledge sharing than other-headed households [57].

In this study, women with a previous CS reported a lower knowledge than those without, indicating a prevalent information gap. Research suggests that CS decisions are doctor-driven primarily on Bangladesh [21], consistent with studies in Iran and India [60, 61]. Women or their families had limited meaningful involvement in decision-making [19] and minimal discussion of risks, benefits, or alternatives, and viewed CS as risk-free in Bangladesh [21, 62]. Additionally, limited physician-patient discussion restricts opportunities for counselling [38] and results in not receiving information during antenatal care [19], which is also consistent with findings from Saudi Arabia [63]. This may explain why many women remain uninformed about CS procedures despite prior CS experience or antenatal care.

The present study also revealed several statistically significant findings associated with positive attitudes toward CS delivery in Bangladesh. The study found that higher monthly family income was associated with more positive attitudes. This is consistent with studies showing that wealthier women in Bangladesh are more likely to undergo CS [32, 43] and that CS rates are higher in private hospitals used by wealthier clients [64]. Similar associations have been reported in Iran [65]. These findings suggest that higher family income may increase access to skilled childbirth services and the likelihood of choosing CS delivery.

The study also found that positive attitudes toward CS delivery decreased as the number of family members increased. Extended family members and traditional norms favoring vaginal birth, together with women’s reliance on family members, may further reduce positive attitudes toward CS [64, 66]. Larger families are also more likely to be poorer and face economic barriers in affording CS. Additionally, provider-related factors, such as inadequate counseling and a focus on financial aspects, may also contribute to lower positive attitudes.

In this study, women in husband-headed households showed lower positive attitudes compared to those in female-headed or other-headed households. This may be explained by the fact that female-headed households are more likely to give birth in health facilities than male-headed households [67, 68]. Similar patterns have been reported in Ethiopia, Gabon, Indonesia, and Sub-Saharan Africa [69–72]. In Bangladesh, where women often have limited decision-making power [73], reduced autonomy in husband-headed households may constrain positive attitudes toward CS delivery.

A prior CS delivery was associated with a substantially lower positive attitude compared to those without a previous CS delivery, consistent with previous studies. This could be linked to most women after a CS, who prefer vaginal birth and are less likely to go through CS again for negative experiences like inadequate information before delivery, economic burden, long recovery time, and social blame [16, 62, 74]. Similar findings have been reported in Ethiopia, the United States, and Japan [75–77]. Postpartum health problems were often left untreated, and women expressed distrust about providers’ motives, including beliefs that unnecessary CS are performed for financial gain [19, 62]. Some women also reported unkind, non-respectful care during labor [19]. Further, the involvement of brokers or “dalal” who transfer women from public to private facilities was documented and linked to extra costs in Bangladesh [16, 19, 42]. These factors may explain why experiencing a CS, with its complications & recovery, was linked to less positive attitudes toward CS.

The study revealed that a higher knowledge scale was related to more positive attitudes. This finding is consistent with previous research showing that limited knowledge can increase fear of vaginal birth and a preference for CS, while improved knowledge of risks and benefits can create a more positive attitude [12]. Although many women had heard of CS, true understanding was low, highlighting the need for better education to prepare and perceptions [47]. Similarly, Nigerian and Pakistani women showed significantly more favorable attitudes toward medically indicated CS, accompanied by greater CS knowledge [78, 79]. This may imply that a lack of knowledge about CS can reduce women’s preparedness or confidence for the procedure.

Overall, the knowledge model explained 21% (Adjusted R square 0.209) variance and the attitude model explained 42.5% (Adjusted R square 0.425) variance, which indicates moderate explanatory power. These findings suggest multi-level strategies and planning that can expand access to quality counselling, strengthen women’s decision-making power, and address socio-economic and healthcare-related factors to reduce unnecessary Caesarean sections.

## Strength & Limitations

The study addresses a clear research gap by using primary data to assess both knowledge and positive attitudes simultaneously toward CS deliveries, focusing on married women of reproductive age in both rural and urban Bangladesh, and provides valuable evidence for targeted maternal health policies and facility-level interventions.

The study has several limitations. Data were obtained from two districts using purposive sampling, and the sample size was relatively small due to time and resource constraints, which may restrict generalizability. The cross-sectional design prevents causal and temporal inference, and social desirability biases may affect self-reported information. The sample included only married women, excluding widowed or divorced women, as well as their husbands’ knowledge and positive attitudes. Lastly, despite demonstrating good reliability (Cronbach’s alpha), several outcomes were measured using newly developed scales that have not been externally validated.

## Conclusion and recommendations

The present study showed the knowledge and positive attitudes toward caesarean section (CS) among married Bangladeshi women and also identified correlates of these outcomes. Our study found that place of residence, household structure, family size, income, and prior CS experience were significantly associated with women’s knowledge and positive attitudes in adjusted analysis. Notably, higher knowledge was associated with more positive attitudes, underscoring the need for well-designed and targeted health education and counselling. Understanding women’s knowledge and positive attitudes toward CS and associated factors is essential for designing targeted, evidence-based policies and interventions that promote safe and appropriate maternal care. These findings suggest that programs and policies should prioritize rural outreach, strengthen provider-patient communication (especially after a CS), ensure high-quality counselling, informed consent practices, and male-inclusive ANC sessions to promote the appropriate use of CS and informed decision-making regarding mode of delivery in Bangladesh. Furthermore, qualitative research should be conducted to better understand the dynamics in this context, highlighting both supply-side and demand-side issues.

## Data Availability

Data cannot be shared publicly because of we are doing another paper based on the data. Data are available from the Chairman of Department of Population Sciences, University of Dhaka (contact via) for researchers who meet the criteria for access to confidential data.

## Acknowledgments

We are grateful to the Department of Population Sciences at the University of Dhaka for permitting us to conduct this research. Lastly, we thank all of the respondents of our study for their valuable support in completing the research.

## Author contributions

**Conceptualization:** Eusrat Jahan, Md. Mahir Faysal.

**Data curation:** Shuvashis Kumar Rimon, Eusrat Jahan.

**Formal analysis:** Shuvashis Kumar Rimon, Eusrat Jahan, Md. Mahir Faysal.

**Investigation:** Eusrat Jahan, Md. Mahir Faysal.

**Methodology:** Shuvashis Kumar Rimon, Eusrat Jahan.

**Supervision:** Md. Mahir Faysal.

**Validation:** Md. Mahir Faysal.

**Writing – original draft:** Shuvashis Kumar Rimon, Eusrat Jahan.

**Writing – review & editing:** Shuvashis Kumar Rimon, Eusrat Jahan, Md. Mahir Faysal.

